# Comorbidities in Early-Onset Sporadic versus Presenilin-1 Mutation-Associated Alzheimer’s Disease Dementia: Evidence for Dependency on Alzheimer’s Disease Neuropathological Changes

**DOI:** 10.1101/2023.08.14.23294081

**Authors:** Diego Sepulveda-Falla, Carlos Andrés Villegas Lanau, Charles White, Geidy E. Serrano, Juliana Acosta-Uribe, Barbara Mejía-Cupajita, Nelson David Villalba-Moreno, Pinzhang Lu, Markus Glatzel, Julia K. Kofler, Bernardino Ghetti, Matthew P. Frosch, Francisco Lopera Restrepo, Kenneth S. Kosik, Thomas G. Beach

## Abstract

Autopsy studies have demonstrated that comorbid neurodegenerative and cerebrovascular disease occur in the great majority of subjects with Alzheimer disease dementia (ADD), and are likely to additively alter the rate of decline or severity of cognitive impairment. The most important of these are Lewy body disease (LBD), TDP-43 proteinopathy and cerebrovascular disease, including white matter rarefaction (WMR) and cerebral infarcts. Comorbidities may interfere with ADD therapeutic trials evaluation of ADD clinical trials as they may not respond to AD-specific molecular therapeutics. It is possible, however, that at least some comorbidities may be, to some degree, secondary consequences of AD pathology, and if this were true then effective AD-specific therapeutics might also reduce the extent or severity of comorbid pathology. Comorbidities in ADD caused by autosomal dominant mutations such as those in the presenilin-1 (*PSEN1*) gene may provide an advantageous perspective on their pathogenesis, and deserve attention because these subjects are increasingly being entered into clinical trials. As ADD associated with *PSEN1* mutations has a presumed single-cause etiology, and the average age at death is under 60, any comorbidities in this setting may be considered as at least partially secondary to the causative AD mechanisms rather than aging, and thus indicate whether effective ADD therapeutics may also be effective for comorbidities. In this study, we sought to compare the rates and types of ADD comorbidities between subjects with early-onset sporadic ADD (EOSADD; subjects dying under age 60) versus ADD associated with different types of *PSEN1* mutations, the most common cause of early-onset autosomal dominant ADD. In particular, we were able to ascertain, for the first time, the prevalences of a fairly complete set of ADD comorbidities in United States (US) *PSEN1* cases as well as the Colombian E280A *PSEN1* kindred. Data for EOSADD and US *PSEN1* subjects (with multiple different mutation types) was obtained from the National Alzheimer Coordinating Center (NACC). Colombian cases all had the E280A mutation and had a set of neuropathological observations classified, like the US cases according to the NACC NP10 definitions. Confirmatory of earlier reports, NACC-defined Alzheimer Disease Neuropathological Changes (ADNC) were consistently very severe in early-onset cases, whether sporadic or in *PSEN1* cases, but were slightly less severe in EOSADD. Amyloid angiopathy was the only AD-associated pathology type with widely-differing severity scores between the 3 groups, with median scores of 3, 2 and 1 in the *PSEN1* Colombia, *PSEN1* US and EOSADD cases, respectively. Apoliprotein E genotype did not show significant proportional group differences for the possession of an E-4 or E-2 allele. Of ADD comorbidities, LBD was most common, being present in more than half of all cases in all 3 groups. For TDP-43 co-pathology, the Colombian *PSEN1* group was the most affected, at about 27%, vs 16% and 11% for the US *PSEN1* and sporadic US cases, respectively. Notably, hippocampal sclerosis and non-AD tau pathological conditions were not present in any of the US or Colombian *PSEN1* cases, and was seen in only 3% of the EOSADD cases. Significant large-vessel atherosclerosis was present in a much larger percentage of Colombian *PSEN1* cases, at almost 20% as compared to 0% and 3% of the US *PSEN1* and EOSADD cases, respectively. Small-vessel disease, or arteriolosclerosis, was much more common than large vessel disease, being present in all groups between 18% and 37%. Gross and microscopic infarcts, however, as well as gross or microscopic hemorrhages, were generally absent or present at very low percentages in all groups. White matter rarefaction (WMR) was remarkably common, at almost 60%, in the US *PSEN1* group, as compared to about 18% in the EOSADD cases, a significant difference. White matter rarefaction was not assessed in the Colombian *PSEN1* cases. The results presented here, as well as other evidence, indicates that LBD, TDP-43 pathology and WMR, as common comorbidities with autosomal dominant and early-onset sporadic ADD, should be considered when planning clinical trials with such subjects as they may increase variability in response rates. However, they may be at least partially dependent on ADNC and thus potentially addressable by anti-amyloid or and/anti-tau therapies.

## Introduction

Autopsy studies have demonstrated that comorbid neurodegenerative and cerebrovascular disease occur in the great majority of subjects with Alzheimer’s disease dementia (ADD), and are likely to additively alter the rate of decline or severity of cognitive impairment [1-5]. Some of these ADD comorbidities including Lewy body disease (LBD), hippocampal sclerosis, TDP-43 proteinopathy and cerebral infarcts, are capable of causing dementia even when ADD neuropathological changes (ADNC) are low or absent. Comorbidities may interfere with the evaluation of ADD clinical trials as they may not respond to ADNC-specific molecular therapeutics. At present, it is not possible to stratify ADD trials for comorbidity presence as there are no proven sensitive and specific diagnostics nor disease-modifying therapies. It is possible, however, that at least some comorbidities may be, to some degree, secondary consequences of ADNC, and if this were true then effective ADNC-specific therapeutics might also reduce the extent or severity of comorbid pathology.

As most reports have focused on the very old, because comorbidities increase with age, it has often been assumed that these would be largely absent in younger, clinical trial-aged sporadic ADD subjects. However, it has recently been shown that comorbidities, particularly LBD, white matter rarefaction (WMR) and TDP-43 proteinopathy, are present even in many cases of early-onset sporadic ADD (EOSADD)[6,7].

Much less is known about the rate of comorbidities in early-onset ADD caused by autosomal dominant mutations in genes such as presenilin-1 (*PSEN1*). This is a critical knowledge gap as ADD in this setting may provide an advantageous perspective on the pathogenesis of ADD comorbidities, and because these subjects are increasingly being entered into clinical trials. It is important, therefore, to understand the comorbidity rates for both early-onset sporadic and inherited ADD so that subject stratification might allow the recognition of significant trial outcomes for subject subsets, even when overall results are negative. As ADD associated with *PSEN1* mutations has a presumed single-cause etiology, and the average age at death is under 60, any comorbidities in this setting may be considered as at least partially secondary to the causative AD mechanisms rather than aging, and thus indicate whether effective ADD therapeutics may also be effective for comorbidities.

In this study, we sought to compare the rates and types of ADD comorbidities between subjects with early-onset sporadic ADD (EOSADD) versus ADD associated with different types of *PSEN1* mutations, the most common cause of early-onset autosomal dominant ADD. In particular, we were able to ascertain, for the first time, the prevalences of a fairly complete set of ADD comorbidities in United States (US) *PSEN1* cases as well as the Colombian E280A *PSEN1* kindred [8-9]. A previous study used NACC data to profile *PSEN1* mutation ADD [10] but less comorbidity data had been reported at that time.

## Methods

The approach used in this study is the same as used in a previous [6] survey of neuropathological comorbidities in sporadic ADD using data from the National Alzheimer Coordinating Center (NACC)[11,12]. Included subject data for sporadic ADD in this study was restricted to those who died under the age of 60 who had dementia and met NIA-AA intermediate or high AD NDNC [13,14], considered to be a sufficient cause of cognitive impairment or dementia; subjects known to have autosomal dominant AD mutations were excluded. Only data from the most recent version of the Neuropathology form (NP10) was used (https://files.alz.washington.edu/documentation/rdd-np.pdf), as this is most inclusive of comorbidity data. Also included were comparative data for *PSEN1* ADD subjects from the United States, derived from the same initial NACC dataset, and from Colombia. The US cases included subjects with several different *PSEN1* mutations while the Colombian cases all had the E280A mutation [8,9]; 36 of the latter have been reported in earlier studies [15,16].

Pathology categories investigated included the major AD-specific lesions: senile or amyloid plaques (NACC variable NPTHAL; all plaque types classified by Thal amyloid phase [17], neurofibrillary tangles (NACC variable NPBRAAK; classified by Braak stage [18,19]), neuritic plaques (NACC variable NPNEUR; classified according to CERAD) [20], diffuse plaques (NACC variable NPDIFF; classified analogously to CERAD neuritic plaques), amyloid angiopathy (NPAMY; classified as none, mild, moderate or severe) and NIA-AA AD Neuropathological Change Level (NACC variable NPADNC [13,14].

Comorbid neurodegenerative conditions investigated included LBD (NACC variable NPLBOD; for this study only presence or absence in any brain region was recorded), TDP-43 pathology (NACC variable NPTDPB; TDP-43 pathology present at least in amygdala, hippocampus or entorhinal area), non-AD tauopathy (NACC variable NPFTDTAU; any non-AD tauopathy including progressive supranuclear palsy, corticobasal degeneration, Pick’s disease, argyrophilic grains, chronic traumatic encephalopathy, or “other”). For each comorbid pathology and ADD group, the proportion of cases possessing that pathology was determined.

Comorbid cerebrovascular conditions investigated included circle of Willis arteriosclerosis (NACC variable NACCAVAS; for this study only “moderate” and “severe” qualified for presence of the condition), old gross cerebral infarcts (NACC variable NPINF; for this study 1 or more large or lacunar infarcts qualified for the condition), old microscopic infarcts (NACC variable NPOLD; for this study 1 or more old microinfarcts qualified for the condition), old microscopic hemorrhages (NACC variable NPOLDD; for this study 1 or more old microhemorrhages qualified for the condition), arteriolosclerosis (NACC variable NACCARTE; for this study only “moderate” and “severe” qualified for presence of the condition) and white matter rarefaction (NACC variable NPWMR; for this study only “moderate” and “severe” qualified for presence of the condition). For each comorbid pathology, the proportion of cases possessing that pathology, relative to those for whom the pathology’s presence or relative absence was specifically recorded, was determined.

Statistical methods used for continuous data included analysis of variance (ANOVA) with post-hoc paired comparisons by the Tukey test. Chi-square and Fisher exact tests were used for comparisons of proportional data. For all tests, the significance level was set at p < 0.05.

## Results

Demographic and mutation data for all cases are shown in Table 1. As a result of our selection criteria, all EOSADD cases were less than 60 years old at death, with a mean age of 56.3 years. For USA *PSEN1* cases the mean age at death was 53.4 years while for Colombian cases it was 57.7 years. For US *PSEN1* subjects a single subject died at age 75; all other US *PSEN1* subjects were 65 or less at death. For Colombian subjects, 4 died at 70 or older. All USA *PSEN1* subjects were Caucasian/white; 13 were non-Hispanic while 6 were Hispanic. There were 12 different *PSEN1* mutations in the US cases while all Colombian cases were multiracial-Hispanic and had the E280A mutation.

**Table 1.** Demographics and mutation genotypes for *PSEN1* and sporadic EOSADD cases.

| Group | Age at Death<br>(mean, SD,<br>range) | Sex | *PSEN1* Mutation Genotypes |  |
| --- | --- | --- | --- | --- |
|  |  |  | Mutation | N (Cases) |
| ***PSEN1* - USA**<br>**N = 19** | 53.4; 8.6; 41-75 | 6 M, 13F | A431E<br>G206A<br>A426P<br>P88L<br>H163R<br>P267A<br>G206E<br>M233L<br>N135S<br>S130L | 5<br>3<br>2<br>2<br>2<br>1<br>1<br>1<br>1<br>1 |
| ***PSEN1* - Colombia**<br>**N = 50** | 57.7; 7.25; 42-79 | 15M, 35F | E280A12 (All) |  |
| **Sporadic - USA**<br>**N = 33** | 56.3; 2.98; 47-59 | 25M; 8 F | N/A |  |

Confirmatory of earlier reports, [6-8,10) ADNC is consistently very severe in early-onset cases, whether they are sporadic or autosomal dominant (Table 2). All 3 groups had median scores that were the highest possible for Thal amyloid phase, Braak neurofibrillary stage, diffuse plaque density and NIA-AA ADNC Level. When the mean scores were compared, however, ANOVA showed significant group differences for Thal amyloid phase, diffuse plaque density and ADNC. On pairwise comparisons for most ADD pathologies, *PSEN1* groups had significantly greater scores than the EOSADD group. Analysis of variance and pairwise comparisons for CERAD neuritic plaque density and Braak neurofibrillary stage were not significant.

**Table 2.** Autopsy data for AD-related neuropathological variables in sporadic EOSADD and *PSEN1* ADD cases. *ApoE*-ε4 and *apoE*-ε2 = number and percentage of cases with one or more *ApoE* ε-4 or ε-2 alleles. Means and medians are shown for the following: NPTHAL = Thal amyloid phase; NPBRAAK = Braak neurofibrillary stage; NPNEUR = CERAD neuritic plaque density; NPDIFF = diffuse plaque density; NPAMY = cerebral amyloid angiopathy density; NPADNC = NIA-AA AD Neuropathological Change level.

| Group | ApoE-4 | ApoE-2 | NPTHAL<sup>1</sup> | NPBRAAK | NPNEUR | NPDIFF<sup>2</sup> | NPADNC<sup>3</sup> | NPAMY<sup>4</sup> |
| --- | --- | --- | --- | --- | --- | --- | --- | --- |
| ***PSEN1*-US** | 6/14<br>42.9% | 2/14<br>14.3% | 5; 5 | 6; 6 | 3; 3 | 3; 3 | 3; 3 | 2.10; 2 |
| ***PSEN1*-COL** | 9/34<br>26.5% | 1/34<br>2.94% | 5; 5 | 6; 6 | 2.9; 3 | 3; 3<br>(n = 36) | 3; 3 | 2.58; 3 |
| **Sporadic-US** | 16/33<br>48.5% | 2/33<br>6.1% | 4.72; 5 | 5.79; 6 | 2.85; 3 | 2.70; 3 | 2.88; 3 | 1.42; 1 |

1. ANOVA $p = 0.002$ ; Tukey $p < 0.05$ for EOSADD vs both *PSEN1* groups. 2. ANOVA $p = 0.022$ ; Tukey $p < 0.05$ for EOSADD vs *PSEN1*-Col group. 3. ANOVA $p = 0.012$ ; Tukey $p < 0.05$ for EOSADD vs *PSEN1* Col group. 4. ANOVA $p = 0.0001$ ; Tukey $p < 0.05$ for EOSADD vs both *PSEN1* groups.

Amyloid angiopathy, which is very consistently present in ADD, was the only AD-associated pathology type with widely-differing severity scores between the 3 groups, with median scores of 3, 2 and 1 in the *PSEN1* Colombia, *PSEN1* US and EOSADD cases, respectively (Table 2). Analysis of variance showed very significant group differences as well as significant differences between both *PSEN1* groups and the EOSADD group.

Apoliporotein E genotype, which strongly influences not only the prevalence and age of onset of ADD but also the prevalence of amyloid angiopathy, did not show significant proportional group differences for the possession of an ε-4 or ε-2 allele, although the US *PSEN1* and EOSADD groups had 1.6-fold and 1.8-fold greater ε-4 possession as compared to the Colombian *PSEN1* group (Table 2). Possession of the ε-2 genotype was relatively rare among all groups, between ∼ 3% and 14%.

The proportion of cases lacking any typical ADD comorbidities, classified here as “AD-Only” cases (Table 3), was low for both of the *PSEN1* groups, less than 30% and only a little higher, 44%, for the EOSADD group. The difference in proportions between groups was not significant.

**Table 3.** Autopsy data for EOSADD and *PSEN1* cases, showing proportions of common ADD comorbidities. Numerator is the number of cases meeting criteria for the comorbidity while denominator is the number of AD cases evaluated for the condition; also given is percentage. AD Only = AD without any of the other conditions in this table and without NPINF, NPOLD and NPOLDD from Table 4; NPLBOD = Lewy body disease; NPHIPSCL = hippocampal sclerosis; TDPNOS = TDP-43 pathology in amygdala, hippocampus or entorhinal area; NPFTDTAU = non-AD tau pathology (PSP, CBD, Pick’s, argyrophilic grains, other).

| Group | AD Only | NPLBOD | NPHIPSCL | TDPNOS<sup>1</sup> | NPFTDTAU |
| --- | --- | --- | --- | --- | --- |
| ***PSEN1*-US** | 2/19; 10.5% | 10/19; 52.6% | 0/19; 0% | 3/19; 15.8% | 0/19; 0% |
| ***PSEN1*-COL** | 5/17; 29.4% | 12/17; 70.6% | 0/50; 0% | 4/15; 26.7% | 0/36; 0% |
| **Sporadic-US** | 8/18; 44.4% | 19/33; 57.6% | 1/32; 3.1% | 2/18; 11.1% | 1/33; 3.0% |

1. Chi-square $p = 0.04$ ; Fisher exact test $p = 0.03$ for sporadic US vs *PSEN1* Col group.

Of ADD comorbidities, LBD was most common (Table 3), being present in more than half of all cases in this study, and was particularly common in the Colombian *PSEN1* and EOSADD cases, at about 70% for each group; the differences in group proportions were not significant, however. Of those cases with Lewy body disease, the brain distribution in the majority of all groups was limbic predominant and amygdala-only, especially in the *PSEN1* groups (Table 4). The brainstem-predominant stage was uncommon in all groups.

**Table 4.** Autopsy data for EOSADD and *PSEN1* cases, showing brain stages of cases with Lewy body disease. Numerator is the number of cases meeting criteria for the stage while denominator is the number of AD cases that were positive in any region for Lewy body disease; also given is percentage. LB-OB = olfactory bulb only; LB-BS = brainstem predominant; LB-Amyg = amygdala predominant; LB-Limb = limbic (transitional); LB-Neo = Neocortical (diffuse).

| Group | LB-OB<sup>1</sup> | LB-BS | LB-Amyg | LB-Limb | LB-Neo |
| --- | --- | --- | --- | --- | --- |
| ***PSEN1*-US** | 3/10; 30% | 0/10; 0% | 3/10; 30% | 4/10; 40% | 0/10; 0% |
| ***PSEN1*-COL** | 1/12; 8.3% | 1/12; 8.3% | 8/12; 66.7% | 4/12; 26.7% | 2/12; 11.8% |
| **Sporadic-US** | 0/33; 0% | 1/33; 3.0% | 10/33; 30.3% | 4/33; 12.1% | 3/33; 10.0% |

1. Fisher exact test $p = 0.002$ for US *PSEN1* vs *PSEN1* Col group.

The olfactory bulb-only stage was not seen in the sporadic group but was significantly more common, at 30%, in the US *PSEN1* group, as compared to 8% for the Colombian *PSEN1* group. The neocortical stage was not seen in the US *PSEN1* group and was only about 10-12% of the EOSADD and Colombian groups; these differences were not significant.

For TDP-43 co-pathology (Table 3), the Colombian *PSEN1* group was the most affected, at about 27%, vs 16% and 11% for the US *PSEN1* and sporadic US cases, respectively. The group differences were significant and the paired comparison between EOSADD and Colombian *PSEN1* cases was significant. For all US *PSEN1* and EOSADD cases, TDP-43 pathology was confined to the amygdala, hippocampus and entorhinal area without any pathology in the neocortex or spinal cord. Colombian *PSEN1* cases were positive in the amygdala with incomplete assessment elsewhere. Notably, hippocampal sclerosis and non-AD tau pathological conditions were not present in any of the US or Colombian *PSEN1* cases, and was seen in only 3% of the EOSADD cases.

Cerebrovascular disease categories varied between the EOSADD and *PSEN1* groups (Table 5). Significant large-vessel atherosclerosis, assessed by NACC as circle of Willis atherosclerosis (NACCAVAS), was present in a much larger percentage of Colombian *PSEN1* cases, at almost 20% as compared to 0% and 3% of the USA *PSEN1* and EOSADD cases, respectively. The group differences were significant as was the comparison between EOSADD cases vs Colombian *PSEN1* cases. Small-vessel disease, or arteriolosclerosis (NACCARTE) was much more common than large vessel disease, being present in all groups between 18% and 37%; the group differences were not significant on a Chi-square test. The consequences of vascular disease include gross and microscopic infarcts, as well as microscopic hemorrhages, but all were generally absent or present at very low percentages in all groups.

**Table 5.** Autopsy data for sporadic EOSADD and *PSEN1* cases, listing major cerebrovascular comorbidities. The numerator is the number of cases meeting criteria for the condition, while the denominator is the number of cases evaluated for the condition. NACCAVAS = severity of atherosclerosis of circle of Willis > mild; NPINF = gross infarcts including lacunes; NPOLD = old microinfarcts; NPOLDD = old cerebral microhemorrhages; NACCARTE = arteriolosclerosis > mild; NPWMR = white matter rarefaction > mild.

| Group | NACCAVAS<sup>1</sup> | NPINF | NPOLD | NPOLDD | NACCARTE | NPWMR<sup>2</sup> |
| --- | --- | --- | --- | --- | --- | --- |
| ***PSEN1*-US** | 0/19; 0% | 2/19; 10.5% | 0/19; 0% | 0/19; 0% | 7/19; 36.8% | 11/19; 57.9% |
| ***PSEN1*-COL** | 9/48; 18.75% | 0/25; 0% | 2/48; 4.2% | 1/48; 2.1% | 9/36; 25% | N/A |
| **Sporadic-US** | 1/33; 3.0% | 0/33; 0% | 2/33; 6.1% | 0/29; 0% | 6/32; 18.7% | 5/28; 17.9% |

1. Chi-square test $p = 0.05$ between groups; Fisher exact test $p = 0.04$ for sporadic vs *PSEN1*-COL.
2. Fisher exact test $p = 0.01$ between EOADD and US *PSEN1* groups.

White matter rarefaction, conventionally regarded as a consequence of arteriolosclerosis, was remarkably common, at almost 60%, in the USA *PSEN1* group, as compared to about 18% in the EOSADD cases, a significant difference. White matter rarefaction was not assessed in the Colombian *PSEN1* cases.

## Discussion

This report updates an earlier comparison by Ringman et al [10] of NACC neuropathology scores for *PSEN1* and sporadic ADD cases (of all ages), based on the prior NP9 NACC neuropathology data elements. We used accumulated data from the NP10 version, which is more extensive in its inclusion of ADD comorbidities. Additionally, we included a comparison with *PSEN1* E280A cases from the Colombian kindred, which have not previously been systematically evaluated for ADD comorbidities.

Confirmatory of earlier NACC-based reports [10,21] we find that the mean severity scores for amyloid plaques, tau-tangles, and especially CAA, are uniformly greater in *PSEN1* ADD cases as compared with those for sporadic ADD, but in this study we have also found that sporadic ADD cases dying before age 60 have amyloid and tau pathology that are almost equivalent in severity to that found in *PSEN1* mutation cases. This concurs with previous reports of sporadic ADD histopathology [6,7] documenting the greatest severity in the cases with earliest onset, and progressively decreased severity with advancing age afterwards. Cerebral amyloid angiopathy (CAA) was the only AD-associated pathology type with markedly differing severity scores between the 3 studied groups, with median scores of 3, 2 and 1 in the *PSEN1* Colombia, *PSEN1* US and EOSADD cases, respectively. The greater CAA severity in *PSEN1* mutation-associated ADD has been previously documented [10] and is of particular relevance because of the greater risk with CAA for amyloid-related imaging abnormalities (ARIA), vasogenic edema and hemorrhage, which are limiting factors for anti-amyloid monoclonal antibody therapy. Other predisposing factors to CAA include the apolipoprotein ε-4 allele, hypercholesterolemia and Hispanic ethnicity [22,23], and, within *PSEN1* cases, mutations beyond codon 200 [10]. The greater prevalence of significant CAA in the E280A Colombian cases might therefore be at least partially due to the higher-codon mutation location, greater proportion with Hispanic ethnicity and greater likelihood of hypercholesterolemia in Colombia as compared to the US [24], although the current set of Colombian subjects had a lower ε-4 carriage rate. Despite the severe CAA severity, only 1/67 combined *PSEN1* cases, and no EOSADD cases, had gross or microscopic brain hemorrhages.

The distribution of apolipoprotein E (*APOE*) ε-4 and ε-2 alleles differed between the studied groups. The ε-4 allele was most common in the *PSEN1* US group, followed by the EOSADD group. Carriage of an *APOE* ε-4 or ε-2 allele has been associated with earlier or later clinical onset, respectively, in both sporadic ADD and *PSEN1* ADD subjects [25,26]. The USA *PSEN1* group had a higher proportion of ε-4 carriers than the Colombian *PSEN1* group, perhaps contributing to their earlier mean onset age in the current study subjects. There was an extremely low proportion (1/34) of Colombian *PSEN1* cases with the ε-2 allele but with this small sample this was not significantly different from the other groups. A single case of the Christchurch *APOE* ε-3 mutation, which in some respects has biochemical similarities with the ε-2 allele, has recently been reported in the Colombian *PSEN1* kindred [27]; this case had a remarkable 30 year delay of clinical onset as well as reduced levels of ADNC [28]. Other disease-modifying genetic loci have been described for *PSEN1* ADD [10,25,29-31] and these offer important clues for the design of disease-modifying strategies.

ADD comorbidities are not restricted to the oldest-old [6,7] as some comorbidity types are common even in early-onset ADD, whether sporadic or inherited. The percentage of cases with ADNC as the sole pathology, defined as the absence of LBD, TDP-43, non-AD tauopathy, infarcts and microhemorrhages, was low in all groups. The NACC *PSEN1* cases had the lowest rate of AD as a sole major pathology, at ∼ 10% while this rate was ∼ 29% for the Colombian *PSEN1* cases and 48% for the EOSADD group.

Of typical ADD comorbidities, LBD is the most common in subjects under age 80 [6], and reaches very high prevalences in this study’s *PSEN1* and EOSADD subjects. Colombian *PSEN1* cases had the highest LBD comorbidity rates, at ∼ 70%, as compared to the US EOSADD and *PSEN1* group at 57% and 53%, respectively. The rates we report here for the *PSEN1* and US sporadic EOSADD cases are considerably higher than the approximately 30% prevalences reported by Ringman et al [10]; this may be due to interim improvements in the immunohistochemical detection of pathological α-synuclein, and to the addition, in the NACC NP10 dataset, of the olfactory bulb-only LBD stage. In another, more recent study of 12 *PSEN1* USA cases [12], 6 (50%) had comorbid α-synuclein pathology.

As the decline in LBD prevalence with age in sporadic ADD closely parallels the decline in AD pathology severity with age [6], and in fact reaches very high prevalences in these *PSEN1* and EOSADD cases, this might suggest that LBD in this setting is largely dependent on AD pathology. The high reported rates of LBD in not only *PSEN1* ADD but also in other forms of autosomal dominant ADD [32-38], Down’s syndrome and even in other, non-Aβ cerebral amyloidoses [39-42], supports this conclusion.

Comorbid TDP-43 proteinopathy has previously been investigated in human *PSEN1* ADD but not in *PSEN1* with the E280A mutation. Two separate USA studies, of a combined 42 cases, both reported limbic-restricted TDP-43 pathology in 17% of subjects with varying *PSEN1* genotypes [42, 43]. We found that TDP-43 pathology is common in *PSEN1* E280A Colombian cases, affecting 27% of subjects, with lesser but still definite rates of 16% and 11% in US *PSEN1* and EOSADD cases. Concurrent TDP-43 proteinopathy is also found in 14% of Down’s syndrome cases [42], further directly implicating ADD pathology. Hippocampal sclerosis, however, which in older people is often associated with hippocampal TDP-43 pathology [44], was present in only 1 EOSADD case and none of the US or Colombian *PSEN1* cases and would therefore seem to require aging as a co-factor. It is likely that non-AD tauopathies, which were rare in these younger subjects, are not common or direct consequences of ADNC and probably are more age-related, as has been shown for sporadic ADD cases with a wide age range [6,7].

Significant large-vessel atherosclerosis, which is usually regarded as age-dependent, was present in a surprisingly large percentage of Colombian *PSEN1* cases, at almost 20% as compared to 0% and 3% of the US *PSEN1* and sporadic US cases, respectively. The Colombian accentuation may be less related to the E280A mutation or ADD pathology than to the generally higher cardiovascular risk factors in Colombia [22] as compared to the USA. Small-vessel disease, or arteriolosclerosis, was more common than large vessel disease in all 3 of the studied groups, with ascending rates between 18% and 37% going from sporadic to Colombian to US *PSEN1* groups. Arteriolosclerosis has been reported to be unrelated to CAA in the *PSEN1* Colombian kindred [16]. Intriguingly, the most severe forms of arteriolosclerosis are found in CADASIL, an autosomal dominant condition [45] caused by Notch-3 mutations, while *PSEN1* acts enzymatically on all Notch genes [46].

The consequences of cerebrovascular disease include gross and microscopic infarcts, as well as gross and microscopic hemorrhages, but all were generally absent or rare in all groups. As mentioned, it might have been expected that microhemorrhages would be much more common in *PSEN1* ADD due to the high rates of CAA. However, these may be underestimated at autopsy as compared to MRI, or when special efforts are made in postmortem examination [47]. In the Alzheimer’s Disease Neuroimaging Initiative, 25% of a mixed group of subjects, with normal cognition, mild cognitive impairment (MCI) and ADD, had microhemorrhages, with a tendency for proportionately more in ADD [48].

White matter rarefaction (WMR), conventionally regarded as a consequence of arteriolosclerosis, was remarkably common, at almost 60%, in the US *PSEN1* group, as compared to 18% in the EOSADD cases. We did not have postmortem estimates of WMR for the Colombian E280A group but previous MR reports [49, 50] found increased mean diffusivity and white matter hyperintensity volumes compared to healthy controls. White matter rarefaction (WMR) has long been noticed to be roughly twice as common in ADD as compared to normal elderly [51-54], and is reportedly pronounced relative to age-matched controls in subjects with *PSEN1* and APP mutations and Down’s syndrome [55-58]. The greater WMR in the US *PSEN1* group studied here, relative to the EOSADD group, may be in part due to the higher proportion of significant arteriolosclerosis, 37% vs 18%, in the *PSEN1* group as compared to the EOSADD group but could also be due to the more severe AD histopathological scores as WMR probably has a dual vascular and AD-related pathogenesis [59-61]. Like LBD, WMR is highly prevalent in autosomal dominant ADD and thus this adds to other evidence that it is at least partially dependent on ADNC. Supporting a vascular contribution in this study both arteriolosclerosis and WMR rarefaction were most common in the NACC *PSEN1* cases.

Effective clinical trials depend on accurate estimates of required subject numbers, which largely depend on accurate assumptions for effect size and variability. Variability of clinical rates of decline will require larger subject numbers to overcome. Effect sizes depend on the efficacy of the therapeutic agent, which may be dependent on a matching of molecular mechanisms of agent and pathogenesis. Neuropathological comorbidities in ADD affect cognition but may have a completely different molecular pathogenesis and therefore might not respond to ADNC-specific therapeutics, leading to loss of efficacy, effect size and probability of clinical trial success. Stratifying ADD subjects by presence and types of accompanying comorbidities might result in increased observed effect sizes in some groups as compared to others, potentially “rescuing” failed clinical trials.

An important limitation of this study is the relatively small subject numbers, especially for the US *PSEN1* group. Some statistical associations may therefore be spurious or undetected.

This study is the first to compare rates of ADD comorbidities in the *PSEN1* Colombian E280A kindred with those in other *PSEN1* mutation types as well as with similarly-aged sporadic ADD cases. The results presented here, as well as other evidence, indicates that LBD, TDP-43 pathology and WMR, as common comorbidities with autosomal dominant and early-onset sporadic ADD, should be considered when planning clinical trials with such subjects as they may increase variability in response rates. However, they may be to some extent at least partially dependent on ADNC and thus potentially addressable by anti-amyloid or anti-tau therapies.

## Data Availability

All data produced in the present study are available upon reasonable request to the authors.

## Acknowledgements

We thank the individuals and the families who participated in this study. We also thank the Grupo de Neurociencias de Antioquia (GNA) staff who helped with the participant recruitment, evaluation and sample processing in Colombia. This study was funded by National Institutes of Health (NIH) grants R01 AG062479-01 (PI Kenneth Kosik, PhD), RF1 NS110048 (PIs Joseph F. Arboleda-Velasquez, PhD/MD, and Diego Sepulveda-Falla, MD), and P01AG025294 (PI William E. Klunk), and by grants to NACC. The NACC database is funded by NIA/NIH Grant U24 AG072122. NACC data are contributed by the NIA-funded ADRCs: P30 AG062429 (PI James Brewer, MD, PhD), P30 AG066468 (PI Oscar Lopez, MD), P30 AG062421 (PI Bradley Hyman, MD, PhD), P30 AG066509 (PI Thomas Grabowski, MD), P30 AG066514 (PI Mary Sano, PhD), P30 AG066530 (PI Helena Chui, MD), P30 AG066507 (PI Marilyn Albert, PhD), P30 AG066444 (PI John Morris, MD), P30 AG066518 (PI Jeffrey Kaye, MD), P30 AG066512 (PI Thomas Wisniewski, MD), P30 AG066462 (PI Scott Small, MD), P30 AG072979 (PI David Wolk, MD), P30 AG072972 (PI Charles DeCarli, MD), P30 AG072976 (PI Andrew Saykin, PsyD), P30 AG072975 (PI David Bennett, MD), P30 AG072978 (PI Neil Kowall, MD), P30 AG072977 (PI Robert Vassar, PhD), P30 AG066519 (PI Frank LaFerla, PhD), P30 AG062677 (PI Ronald Petersen, MD, PhD), P30 AG079280 (PI Eric Reiman, MD), P30 AG062422 (PI Gil Rabinovici, MD), P30 AG066511 (PI Allan Levey, MD, PhD), P30 AG072946 (PI Linda Van Eldik, PhD), P30 AG062715 (PI Sanjay Asthana, MD, FRCP), P30 AG072973 (PI Russell Swerdlow, MD), P30 AG066506 (PI Todd Golde, MD, PhD), P30 AG066508 (PI Stephen Strittmatter, MD, PhD), P30 AG066515 (PI Victor Henderson, MD, MS), P30 AG072947 (PI Suzanne Craft, PhD), P30 AG072931 (PI Henry Paulson, MD, PhD), P30 AG066546 (PI Sudha Seshadri, MD), P20 AG068024 (PI Erik Roberson, MD, PhD), P20 AG068053 (PI Justin Miller, PhD), P20 AG068077 (PI Gary Rosenberg, MD), P20 AG068082 (PI Angela Jefferson, PhD), P30 AG072958 (PI Heather Whitson, MD), P30 AG072959 (PI James Leverenz, MD).

## Notes

### Competing Interest Statement

Thomas G. Beach is a paid consultant for Aprinoia Therapeutics and is a scientific advisory board member for Vivid Genomics.

### Author Declarations

The ethics committee/IRB of the University of Antioquia, Medellin, Colombia gave ethical approval for this work. The ethics committee/IRB of Banner Sun Health Research Institute gave ethical approval for this work.

## References

[1] Boyle PA, Yu L, Leurgans SE, Wilson RS, Brookmeyer R, Schneider JA, Bennett DA (2019) Attributable risk of Alzheimer’s dementia attributed to age-related neuropathologies. Ann Neurol 85, 114–124.

[2] Jansen WJ, Wilson RS, Visser PJ, Nag S, Schneider JA, James BD, Leurgans SE, Capuano AW, Bennett DA, Boyle PA (2018) Age and the association of dementia-related pathology with trajectories of cognitive decline. Neurobiol Aging 61, 138–145.

[3] Malek-Ahmadi M, Beach TG, Zamrini E, Adler CH, Sabbagh MN, Shill HA, Jacobson SA, Belden CM, Caselli RJ, Woodruff BK, Rapscak SZ, Ahern GL, Shi J, Caviness JN, Driver-Dunckley E, Mehta SH, Shprecher DR, Spann BM, Tariot P, Davis KJ, Long KE, Nicholson LR, Intorcia A, Glass MJ, Walker JE, Callan M, Curry J, Cutler B, Oliver J, Arce R, Walker DG, Lue LF, Serrano GE, Sue LI, Chen K, Reiman EM (2019) Faster cognitive decline in dementia due to Alzheimer disease with clinically undiagnosed Lewy body disease. PLoS One 14, e0217566.

[4] Caselli RJ, Beach TG, Knopman DS, Graff-Radford NR (2017) Alzheimer Disease: Scientific Breakthroughs and Translational Challenges. Mayo Clin Proc 92, 978–994.

[5] Beach TG, Adler CH, Sue LI, Serrano G, Shill HA, Walker DG, Lue L, Roher AE, Dugger BN, Maarouf C, Birdsill AC, Intorcia A, Saxon-Labelle M, Pullen J, Scroggins A, Filon J, Scott S, Hoffman B, Garcia A, Caviness JN, Hentz JG, Driver-Dunckley E, Jacobson SA, Davis KJ, Belden CM, Long KE, Malek-Ahmadi M, Powell JJ, Gale LD, Nicholson LR, Caselli RJ, Woodruff BK, Rapscak SZ, Ahern GL, Shi J, Burke AD, Reiman EM, Sabbagh MN (2015) Arizona Study of Aging and Neurodegenerative Disorders and Brain and Body Donation Program. Neuropathology 35, 354–389.

[6] Beach TG, Malek-Ahmadi M (2021) Alzheimer’s disease neuropathological comorbidities are common in the younger-old. J Alzheimer’s Dis 79, 389–400.

[7] Spina S, La Joie R, Petersen C, Nolan AL, Cuevas D, Cosme C, Hepker M, Hwang JH, Miller ZA, Huang EJ, Karydas AM, Grant H, Boxer AL, Gorno-Tempini ML, Rosen HJ, Kramer JH, Miller BL, Seeley WW, Rabinovici GD, Grinberg LT (2021) Comorbid neuropathological diagnoses in early versus late-onset Alzheimer’s disease. Brain 144, 2186–2198.

[8] Sepulveda-Falla D, Glatzel M, Lopera F (2012) Phenotypic profile of early-onset familial Alzheimer’s disease caused by presenilin-1 E280A mutation. J Alzheimer’s Dis 32, 1–12.

[9] Fuller JT, Cronin-Golomb A, Gatchel JR, Norton DJ, Guzmán-Vélez E, Jacobs HIL, Hanseeuw B, Pardilla-Delgado E, Artola A, Baena A, Bocanegra Y, Kosik KS, Chen K, Tariot PN, Johnson K, Sperling RA, Reiman EM, Lopera F, Quiroz YT (2019) Biological and cognitive markers of presenilin1 E280A autosomal dominant Alzheimer’s disease: a comprehensive review of the Colombian kindred. J Prev Alzheimers Dis 6, 112–120.

[10] Ringman JM, Monsell S, Ng DW, Zhou Y, Nguyen A, Coppola G, Van B, V, Mendez MF, Tung S, Weintraub S, Mesulam MM, Bigio EH, Gitelman DR, Fisher-Hubbard AO, Albin RL, Vinters HV (2016) Neuropathology of Autosomal Dominant Alzheimer Disease in the National Alzheimer Coordinating Center Database. J Neuropathol Exp Neurol 75, 284–290.

[11] Besser L, Kukull W, Knopman DS, Chui H, Galasko D, Weintraub S, Jicha G, Carlsson C, Burns J, Quinn J, Sweet RA, Rascovsky K, Teylan M, Beekly D, Thomas G, Bollenbeck M, Monsell S, Mock C, Zhou XH, Thomas N, Robichaud E, Dean M, Hubbard J, Jacka M, Schwabe-Fry K, Wu J, Phelps C, Morris JC (2018) Version 3 of the National Alzheimer’s Coordinating Center’s Uniform Data Set. Alzheimer Dis Assoc Disord 32, 351–358.

[12] Besser LM, Kukull WA, Teylan MA, Bigio EH, Cairns NJ, Kofler JK, Montine TJ, Schneider JA, Nelson PT (2018) The Revised National Alzheimer’s Coordinating Center’s Neuropathology Form-Available Data and New Analyses. J Neuropathol Exp Neurol 77, 717–726.

[13] Hyman BT, Phelps CH, Beach TG, Bigio EH, !Lost Data, Carrillo MC, Dickson DW, Duyckaerts C, Frosch MP, Masliah E, Mirra SS, Nelson PT, Schneider JA, Thal DR, Thies B, Trojanowski JQ, Vinters HV, Montine TJ (2012) National Institute on Aging-Alzheimer’s Association guidelines for the neuropathologic assessment of Alzheimer’s disease. Alzheimers Dement 8, 1–13.

[14] Montine TJ, Phelps CH, Beach TG, Bigio EH, Cairns NJ, Dickson DW, Duyckaerts C, Frosch MP, Masliah E, Mirra SS, Nelson PT, Schneider JA, Thal DR, Trojanowski JQ, Vinters HV, Hyman BT (2012) National Institute on Aging-Alzheimer’s Association guidelines for the neuropathologic assessment of Alzheimer’s disease: a practical approach. Acta Neuropathol 123, 1–11.

[15] Sepulveda-Falla D, Chavez-Gutierrez L, Portelius E, Vélez JI, Dujardin S, Barrera-Ocampo A, Dinkel F, Hagel C, Puig B, Mastronardi C, Lopera F, Hyman BT, Blennow K, Arcos-Burgos M, de Strooper B, Glatzel M. A multifactorial model of pathology for age of onset heterogeneity in familial Alzheimer’s disease. Acta Neuropathol. 2021 Feb;141(2):217–233.

[16] Littau JL, Velilla L, Hase Y, Villalba-Moreno ND, Hagel C, Drexler D, Osorio Restrepo S, Villegas A, Lopera F, Vargas S, Glatzel M, Krasemann S, Quiroz YT, Arboleda-Velasquez JF, Kalaria R, Sepulveda-Falla D (2022) Evidence of beta-amyloid independent small vessel disease in familial Alzheimer’s disease. Brain Pathol 32, e13097.

[17] Thal DR, Rub U, Orantes M, Braak H (2002) Phases of A beta-deposition in the human brain and its relevance for the development of AD. Neurology 58, 1791–1800.

[18] Alafuzoff I, Arzberger T, Al-Sarraj S, Bodi I, Bogdanovic N, Braak H, Bugiani O, Del-Tredici K, Ferrer I, Gelpi E, Giaccone G, Graeber MB, Ince P, Kamphorst W, King A, Korkolopoulou P, Kovacs GG, Larionov S, Meyronet D, Monoranu C, Parchi P, Patsouris E, Roggendorf W, Seilhean D, Tagliavini F, Stadelmann C, Streichenberger N, Thal DR, Wharton SB, Kretzschmar H (2008) Staging of neurofibrillary pathology in Alzheimer’s disease: a study of the BrainNet Europe Consortium. Brain Pathol 18, 484–496.

[19] Braak H, Alafuzoff I, Arzberger T, Kretzschmar H, Del TK (2006) Staging of Alzheimer disease-associated neurofibrillary pathology using paraffin sections and immunocytochemistry. Acta Neuropathol 112, 389–404.

[20] Mirra SS, Heyman A, McKeel D, Sumi SM, Crain BJ, Brownlee LM, Vogel FS, Hughes JP, van Belle G, Berg L (1991) The Consortium to Establish a Registry for Alzheimer’s Disease (CERAD). Part II. Standardization of the neuropathologic assessment of Alzheimer’s disease. Neurology 41, 479–486.

[21] Gómez-Isla T, Growdon WB, McNamara MJ, Nochlin D, Bird TD, Arango JC, Lopera F, Kosik KS, Lantos PL, Cairns NJ, Hyman BT (1999). The impact of different presenilin 1 and presenilin 2 mutations on amyloid deposition, neurofibrillary changes and neuronal loss in the familial Alzheimer’s disease brain: evidence for other phenotype-modifying factors. Brain 122, 1709–19.

[22] Ringman JM, Sachs MC, Zhou Y, Monsell SE, Saver JL, Vinters HV (2014) Clinical predictors of severe cerebral amyloid angiopathy and influence of APOE genotype in persons with pathologically verified Alzheimer disease. JAMA Neurol 71, 878–83.

[23] Xu C, Apostolova LG, Oblak AL, Gao S (2020) Association of hypercholesterolemia with Alzheimer’s disease pathology and cerebral amyloid angiopathy. J Alzheimer’s Dis 73, 1305–1311.

[24] Miranda JJ, Herrera VM, Chirinos JA, Gómez LF, Perel P, Pichardo R, González A, Sánchez JR, Ferreccio C, Aguilera X, Silva E, Oróstegui M, Medina-Lezama J, Pérez CM, Suárez E, Ortiz AP, Rosero L, Schapochnik N, Ortiz Z, Ferrante D, Casas JP, Bautista LE (2013) Major cardiovascular risk factors in Latin America: a comparison with the United States. The Latin American Consortium of Studies in Obesity (LASO). PLoS One 8, e54056.

[25] Vélez JI, Lopera F, Sepulveda-Falla D, Patel HR, Johar AS, Chuah A, Tobón C, Rivera D, Villegas A, Cai Y, Peng K, Arkell R, Castellanos FX, Andrews SJ, Silva Lara MF, Creagh PK, Easteal S, de Leon J, Wong ML, Licinio J, Mastronardi CA, Arcos-Burgos M (2016) APOE-E2 allele delays age of onset in PSEN1 E280A Alzheimer’s disease. Mol Psychiatry 21, 916–24.

[26] Cacace R, Sleegers K, Van Broeckhoven C (2016). Molecular genetics of early-onset Alzheimer’s disease revisited. Alzheimers Dement 12, 733–48.

[27] Arboleda-Velasquez JF, Lopera F, O’Hare M, Delgado-Tirado S, Marino C, Chmielewska N, Saez-Torres KL, Amarnani D, Schultz AP, Sperling RA, Leyton-Cifuentes D, Chen K, Baena A, Aguillon D, Rios-Romenets S, Giraldo M, Guzmán-Vélez E, Norton DJ, Pardilla-Delgado E, Artola A, Sanchez JS, Acosta-Uribe J, Lalli M, Kosik KS, Huentelman MJ, Zetterberg H, Blennow K, Reiman RA, Luo J, Chen Y, Thiyyagura P, Su Y, Jun GR, Naymik M, Gai X, Bootwalla M, Ji J, Shen L, Miller JB, Kim LA, Tariot PN, Johnson KA, Reiman EM, Quiroz YT (2019). Resistance to autosomal dominant Alzheimer’s disease in an APOE3 Christchurch homozygote: a case report. Nat Med 25, 1680–1683.

[28] Sepulveda-Falla D, Sanchez JS, Almeida MC, Boassa D, Acosta-Uribe J, Vila-Castelar C, Ramirez-Gomez L, Baena A, Aguillon D, Villalba-Moreno ND, Littau JL, Villegas A, Beach TG, White CL 3rd, Ellisman M, Krasemann S, Glatzel M, Johnson KA, Sperling RA, Reiman EM, Arboleda-Velasquez JF, Kosik KS, Lopera F, Quiroz YT (2022) Distinct tau neuropathology and cellular profiles of an APOE3 Christchurch homozygote protected against autosomal dominant Alzheimer’s dementia. Acta Neuropathol 144, 589–601

[29] Sepulveda-Falla D (2023) Resistant and resilient mutations in protection against familial Alzheimer’s disease: learning from nature. Mol Neurodegener 18, 36.

[30] Lopera F, Marino C, Chandrahas AS, O’Hare M, Villalba-Moreno ND, Aguillon D, Baena A, Sanchez JS, Vila-Castelar C, Ramirez Gomez L, Chmielewska N, Oliveira GM, Littau JL, Hartmann K, Park K, Krasemann S, Glatzel M, Schoemaker D, Gonzalez-Buendia L, Delgado-Tirado S, Arevalo-Alquichire S, Saez-Torres KL, Amarnani D, Kim LA, Mazzarino RC, Gordon H, Bocanegra Y, Villegas A, Gai X, Bootwalla M, Ji J, Shen L, Kosik KS, Su Y, Chen Y, Schultz A, Sperling RA, Johnson K, Reiman EM, Sepulveda-Falla D, Arboleda-Velasquez JF, Quiroz YT (2023) Resilience to autosomal dominant Alzheimer’s disease in a Reelin-COLBOS heterozygous man. Nat Med 29, 1243–1252.

[31] Cochran JN, Acosta-Uribe J, Madrigal L, Aguillón D, Garcia GP, Giraldo MM, Taylor JW, Bradley J, Fulton-Howard B, Andrews SJ, Acosta-Baena N, Alzate D, Garcia GP, Piedrahita F, Lopez HE, Anderson AG, Rodriguez-Nunez I, Roberts K, Absher D, Myers RM, Beecham GW, Reitz C, Rizzardi LF, Fernandez MV, Goate AM, Cruchaga C. Renton AE, Lopera F, Kosik KS (2020) Genetic Associations with Age at Dementia Onset in the PSEN1 E280A Colombian Kindred. medRxiv, doi: https://doi.org/10.1101/2020.09.23.20198424.

[32] Halliday G, Brooks W, Arthur H, Creasey H, Broe GA (1997) Further evidence for an association between a mutation in the APP gene and Lewy body formation. Neurosci Lett 227, 49–52.

[33] Ishikawa A, Piao YS, Miyashita A, Kuwano R, Onodera O, Ohtake H, Suzuki M, Nishizawa M, Takahashi H (2005) A mutant PSEN1 causes dementia with Lewy bodies and variant Alzheimer’s disease. Ann Neurol 57, 429–434.

[34] Rosenberg CK, Pericak-Vance MA, Saunders AM, Gilbert JR, Gaskell PC, Hulette CM (2000) Lewy body and Alzheimer pathology in a family with the amyloid-beta precursor protein APP717 gene mutation. Acta Neuropathol 100, 145–152.

[35] Lantos PL, Ovenstone IM, Johnson J, Clelland CA, Roques P, Rossor MN (1994) Lewy bodies in the brain of two members of a family with the 717 (Val to Ile) mutation of the amyloid precursor protein gene. Neurosci Lett 172, 77–79.

[36] Lippa CF, Fujiwara H, Mann DM, Giasson B, Baba M, Schmidt ML, Nee LE, O’Connell B, Pollen DA, St George-Hyslop P, Ghetti B, Nochlin D, Bird TD, Cairns NJ, Lee VM, Iwatsubo T, Trojanowski JQ (1998) Lewy bodies contain altered alpha-synuclein in brains of many familial Alzheimer’s disease patients with mutations in presenilin and amyloid precursor protein genes. Am J Pathol 153, 1365–1370.

[37] Leverenz JB, Fishel MA, Peskind ER, Montine TJ, Nochlin D, Steinbart E, Raskind MA, Schellenberg GD, Bird TD, Tsuang D (2006) Lewy body pathology in familial Alzheimer disease: evidence for disease-and mutation-specific pathologic phenotype. Arch Neurol 63, 370–376.

[38] Kaneko H, Kakita A, Kasuga K, Nozaki H, Ishikawa A, Miyashita A, Kuwano R, Ito G, Iwatsubo T, Takahashi H, Nishizawa M, Onodera O, Sisodia SS, Ikeuchi T (2007) Enhanced accumulation of phosphorylated alpha-synuclein and elevated beta-amyloid 42/40 ratio caused by expression of the presenilin-1 deltaT440 mutant associated with familial Lewy body disease and variant Alzheimer’s disease. J Neurosci 27, 13092–13097.

[39] Haltia M, Ghiso J, Wisniewski T, Kiuru S, Miller D, Frangione B (1991) Gelsolin variant and beta-amyloid co-occur in a case of Alzheimer’s with Lewy bodies. Neurobiol Aging 12, 313–316.

[40] Bugiani O, Giaccone G, Piccardo P, Morbin M, Tagliavini F, Ghetti B (2000) Neuropathology of Gerstmann-Straussler-Scheinker disease. Microsc Res Tech 50, 10–15.

[41] Sutovsky S, Smolek T, Turcani P, Petrovic R, Brandoburova P, Jadhav S, Novak P, Attems J, Zilka N (2018). Neuropathology and biochemistry of early onset familial Alzheimer’s disease caused by presenilin-1 missense mutation Thr116Asn. J Neural Transm (Vienna)125, 965–976.

[42] Lippa CF, Rosso AL, Stutzbach LD, Neumann M, Lee VM, Trojanowski JQ. Transactive response DNA-binding protein 43 burden in familial Alzheimer disease and Down syndrome. Arch Neurol. 2009 Dec;66(12):1483–8.

[43] Abrahamson EE, Kofler JK, Becker CR, Price JC, Newell KL, Ghetti B, Murrell JR, McLean CA, Lopez OL, Mathis CA, Klunk WE, Villemagne VL, Ikonomovic MD (2022) 11C-PiB PET can underestimate brain amyloid-β burden when cotton wool plaques are numerous. Brain 145:2161–2176.

[44] Gauthreaux KM, Teylan MA, Katsumata Y, Mock C, Culhane JE, Chen YC, Chan KCG, Fardo DW, Dugan AJ, Cykowski MD, Jicha GA, Kukull WA, Nelson PT (2022) Limbic-Predominant Age-Related TDP-43 Encephalopathy: Medical and Pathologic Factors Associated With Comorbid Hippocampal Sclerosis. Neurology 98, e1422–e1433.

[45] Fang C, Magaki SD, Kim RC, Kalaria RN, Vinters HV, Fisher M (2023) Arteriolar neuropathology in cerebral microvascular disease. Neuropathol Appl Neurobiol 49, e12875.

[46] Chávez-Gutiérrez L, Bammens L, Benilova I, Vandersteen A, Benurwar M, Borgers M, Lismont S, Zhou L, Van Cleynenbreugel S, Esselmann H, Wiltfang J, Serneels L, Karran E, Gijsen H, Schymkowitz J, Rousseau F, Broersen K, De Strooper B (2012) The mechanism of γ-Secretase dysfunction in familial Alzheimer disease. EMBO J 31, 2261–74.

[47] Tanskanen M, Makela M, Myllykangas L, Rastas S, Sulkava R, Paetau A (2012) Intracerebral hemorrhage in the oldest old: a population-based study (vantaa 85+). Front Neurol 3, 103–

[48] Kantarci K, Gunter JL, Tosakulwong N, Weigand SD, Senjem MS, Petersen RC, Aisen PS, Jagust WJ, Weiner MW, Jack CR, Jr. (2013) Focal hemosiderin deposits and beta-amyloid load in the ADNI cohort. Alzheimers Dement 9, S116–S123.

[49] Schoemaker D, Zanon Zotin MC, Chen K, Igwe KC, Vila-Castelar C, Martinez J, Baena A, Fox-Fuller JT, Lopera F, Reiman EM, Brickman AM, Quiroz YT (2022) White matter hyperintensities are a prominent feature of autosomal dominant Alzheimer’s disease that emerge prior to dementia. Alzheimers Res Ther 14, 89.

[50] Parra MA, Saarimäki H, Bastin ME, Londoño AC, Pettit L, Lopera F, Della Sala S, Abrahams S (2015) Memory binding and white matter integrity in familial Alzheimer’s disease. Brain 138, 1355–69.

[51] Brun A, Englund E (1986) A white matter disorder in dementia of the Alzheimer type: a pathoanatomical study. Ann Neurol 19, 253–262.

[52] Englund E, Brun A, Alling C (1988) White matter changes in dementia of Alzheimer’s type. Biochemical and neuropathological correlates. Brain 111, 1425–1439.

[53] Mirsen TR, Lee DH, Wong CJ, Diaz JF, Fox AJ, Hachinski VC, Merskey H (1991) Clinical correlates of white-matter changes on magnetic resonance imaging scans of the brain. Arch Neurol 48, 1015–1021.

[54] Prins ND, Scheltens P (2015) White matter hyperintensities, cognitive impairment and dementia: an update. Nat Rev Neurol 11, 157–165.

[55] Li X, Westman E, Ståhlbom AK, Thordardottir S, Almkvist O, Blennow K, Wahlund LO, Graff C (2015) White matter changes in familial Alzheimer’s disease. J Intern Med 278, 211–8.

[56] Rosas HD, Hsu E, Mercaldo ND, Lai F, Pulsifer M, Keator D, Brickman AM, Price J, Yassa M, Hom C, Krinsky-McHale SJ, Silverman W, Lott I, Schupf N (2020) Alzheimer-related altered white matter microstructural integrity in Down syndrome: A model for sporadic AD? Alzheimers Dement (Amst) 12, e12040.

[57] Lee S, Viqar F, Zimmerman ME, Narkhede A, Tosto G, Benzinger TL, Marcus DS, Fagan AM, Goate A, Fox NC (2016) White matter hyperintensities are a core feature of Alzheimer’s disease: evidence from the dominantly inherited Alzheimer network. Annals of Neurology 79, 929–939.

[58] Roher AE, Maarouf CL, Malek-Ahmadi M, Wilson J, Kokjohn TA, Daugs ID, Whiteside CM, Kalback WM, Macias MP, Jacobson SA, Sabbagh MN, Ghetti B, Beach TG (2013) Subjects harboring presenilin familial Alzheimer’s disease mutations exhibit diverse white matter biochemistry alterations. Am J Neurodegener Dis 2, 187–207.

[59] McAleese KE, Miah M, Graham S, Hadfield GM, Walker L, Johnson M, Colloby SJ, Thomas AJ, DeCarli C, Koss D, Attems J (2021) Frontal white matter lesions in Alzheimer’s disease are associated with both small vessel disease and AD-associated cortical pathology. Acta Neuropathol 142, 937–950.

[60] Blevins BL, Vinters HV, Love S, Wilcock DM, Grinberg LT, Schneider JA, Kalaria RN, Katsumata Y, Gold BT, Wang DJJ, Ma SJ, Shade LMP, Fardo DW, Hartz AMS, Jicha GA, Nelson KB, Magaki SD, Schmitt FA, Teylan MA, Ighodaro ET, Phe P, Abner EL, Cykowski MD, Van Eldik LJ, Nelson PT (2021) Brain arteriolosclerosis. Acta Neuropathol 141, 1–24.

[61] Beach TG, Sue LI, Scott S, Intorcia AJ, Walker JE, Arce RA, Glass MJ, Borja CI, Cline MP, Hemmingsen SJ, Qiji S, Stewart A, Martinez KN, Krupp A, McHattie R, Mariner M, Lorenzini I, Kuramoto A, Long KE, Tremblay C, Caselli RJ, Woodruff BK, Rapscak SZ, Belden CM, Goldfarb D, Choudhury P, Driver-Dunckley ED, Mehta SH, Sabbagh MN, Shill HA, Atri A, Adler CH, Serrano GE. Cerebral white matter rarefaction has both neurodegenerative and vascular causes and may primarily be a distal axonopathy (2023) J Neuropathol Exp Neurol 82, 457–466.

